# Causal analyses using education-health linked data for England: a case study

**DOI:** 10.64898/2026.03.13.26348340

**Authors:** Bianca L De Stavola, Andrea Aparicio Castro, Vincent G. Nguyen, Kate Lewis, Lorraine Dearden, Katie Harron, Ania Zylbersztejn, Julia Shumway, Ruth Gilbert, the HOPE study team

## Abstract

**Introduction:** This article summarises lessons learnt from the Health Outcomes for young People throughout Education (HOPE) Study and serves as a real world, transferable application for addressing causal questions using administrative data. The HOPE study applied causal methods to analyses of administrative data in Education and Child Health Insights from Linked Data (ECHILD) aimed at studying the effectiveness of provision for special educational needs and disability (SEND) on health and education outcomes.

**Methods:** Defining causal questions regarding the impact of SEND provision required judicious mapping of the question onto the data, leading to the selection of appropriate measures of effect, transparent handling of the data and control of confounding factors to estimate effects. We adopted the target trial emulation framework to guide these steps. Having encountered specific computational challenges in estimating the effects of interest, we simulated data that resembled the HOPE study and used them to practice the implementation of alternative estimation methods and to study impact of some of their assumptions.

**Results:** The creation and analysis of the simulated data provided valuable insights. First, we learned the importance of aligning the target of estimation with the causal question at hand. Second, we observed how deviations from assumptions specific to each estimation method can affect results. Third, we highlighted the benefits of employing alternative estimation methods as sensitivity tools that can aid the interpretation of the resulting estimates. Finally, we offer user-friendly code in two programming languages (R and Stata) and accompanying simulated data to facilitate the implementation of these methods for similar causal questions.

**Conclusion:** We recommend users of administrative data to fully specify -and possibly revise- the causal questions they wish to address and to carefully examine and compare assumptions, implementation and results obtained using alternative estimation methods.

## Introduction

Policymakers require evidence on the effectiveness of both new and existing interventions. The growing accessibility of large-scale administrative data presents a valuable opportunity to generate this evidence. However, the use of such data for causal inference demands attention not only towards issues of data quality [1], but also towards the formulation of the target of investigation and the extraction of the relevant data. We share our lessons learnt in investigating special educational needs and disability (SEND) provision in England to demonstrate some of these additional challenges. The work was part of the HOPE (Health Outcomes of young People throughout Education) study and the data source was ECHILD [2] (full details in Gilbert et al 2026 [3]).

### Case study

One of HOPE’s main aims was to study the impact of SEND provision on health and education outcomes during primary education. As this is a causal investigation, we addressed it following what is known as the “causal roadmap” [4, 5, 6]). This consists of:

i. *Stating the causal question*
ii. *Articulating its scope*
iii. *Translating the question into a statistic that captures the effect of interest*
iv. *Explicit setting out of assumptions for its estimation*
v. *Estimating and critically interpretating the results*

The first two steps required careful mapping of the question to the available the data, while the latter three call for adoption of language and methods from modern causal inference. We adopted the target trial emulation (TTE) framework [7] to perform these steps. TTE consists of first conceptualising an *ideal target trial* that would address the causal investigation of interest, before determining how to emulate this trial using the available observational data. TTE compels researchers to define the study’s eligibility criteria and the timing of potential interventions and follow-ups in relation to the data at hand (step *(ii)*). It also highlights the importance of linking causal question and target of estimation (step *(iii)*) and to perform sensitivity analyses by comparing results obtained using alternative estimation methods (*(iv)* and *(v)*). The adoption of this approach led us to refine the initial, vaguely defined, causal question (“*What is the impact of SEND provision on health and education outcomes?*”) and to pay greater attention to how the administrative data were handled. Furthermore, we used multiple estimation methods, developing bespoke code as software capable of estimating causal effects in our scenarios was not readily available. We checked our code by testing it on simulated data that mirrored key features of ECHILD.

## Methods

### The causal roadmap

Below we review the challenges we encountered in applying the five steps of the causal roadmap implemented using the TTE framework (Figure 1).

**Figure 1.**
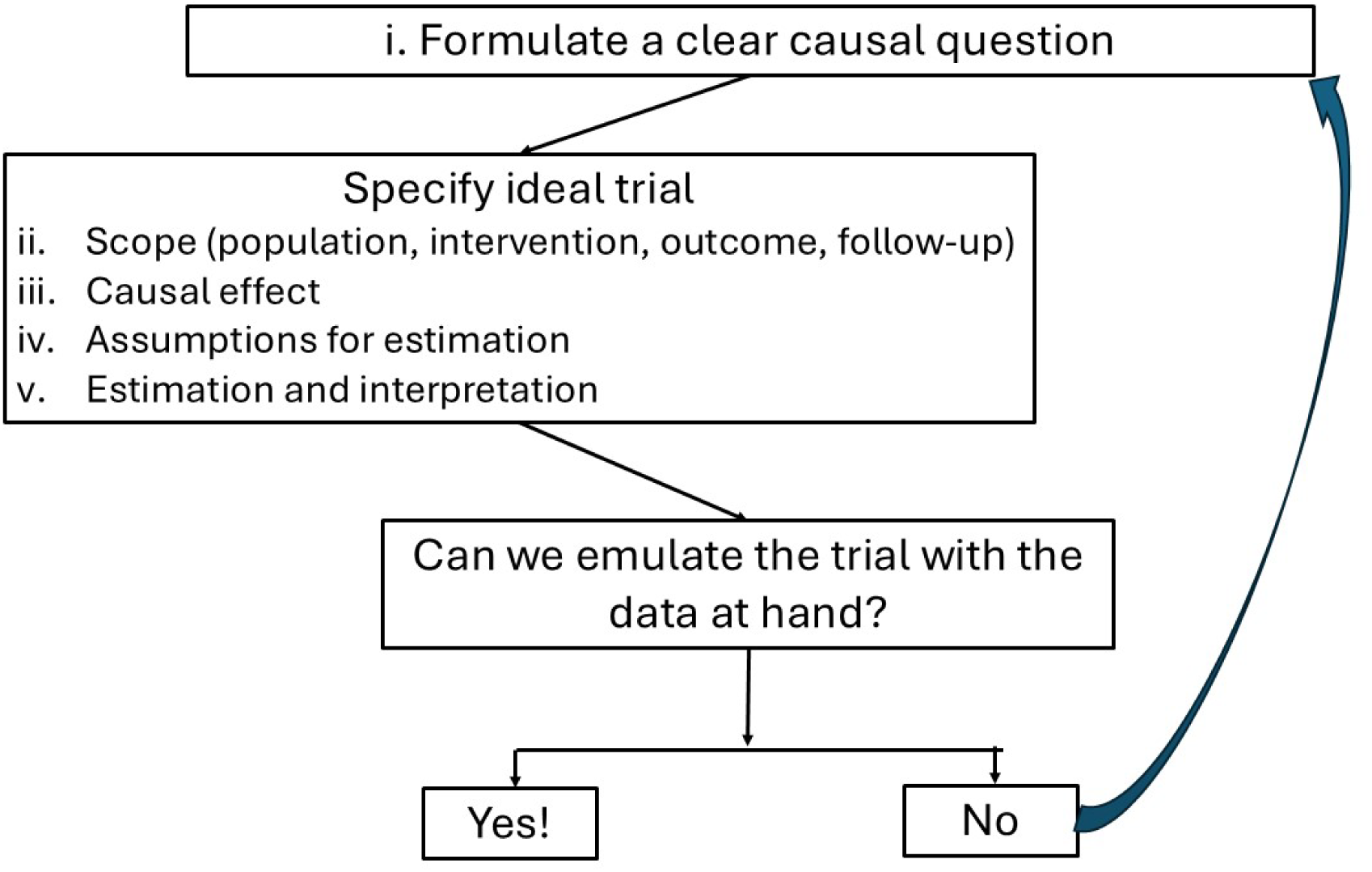
– Target trial emulation implementation of the “causal roadmap”

#### i. The causal question

The broad causal question addressed by the HOPE study was too loosely formulated to be conceptualised via a randomised controlled trial, indeed it was not “well-defined” [8, 9]. For example, it did not clarify when the intervention should occur (“*Just in Year 1 or sustained over the entire period of primary education?)*, nor the timeframe for the outcomes (“*Up to the end of the year following intervention or over the entire primary school period?”*). It also concerned three [3]. Using the TTE framework as guidance, we therefore expanded the original causal question into multiple questions for each outcome [10, 11, 12]. Here we focus on one exemplar outcome: the number of unauthorised school absences (hereafter UA) up to the end of primary education (Year 6). The new questions were:

*Q1.* Does SEND provision in Year 1 affect the number of UA from Year 2 to Year 6 *(long-term effect*)?
*Q2. Does SEND provision in Year t affect the number of UA in Year t +* 1 *(shortterm effect)?* Does SEND provision in Year *t* affect the number of UA in Year t+ 1 *(short-term effect*)?
*Q3.* Does *sustained SEND provision* in Year 1, 2 and 3 affect the number of UA from Year 2 to Year 6?

#### ii. The scope

Articulating the scope of these questions requires first identifying the population(s) for which they are most relevant. Being aware that SEND provision is most likely to have an impact on children in need of such support, our populations of interest became children with specific health conditions for whom SEND provision is potentially beneficial, as SEND is not expected to be suitable for the wider population. As a first step, we focussed on two such groups: children with cleft lip and/or palate (CLP) [10] and children with cerebral palsy (CP) [11]. Within these groups, however, children with additional major abnormalities are more likely to have more pronounced needs while the available information within ECHILD is too coarse to permit proper adjustment for these differentials. The original questions were therefore redefined (following the loop from “No” to *“i.”* in Figure 1) to concern only children with no other major abnormalities (N≈10,000).

Intervention, follow-up and outcome varied across the three questions. Q1 refers to an intervention in Year 1 with number of UA assessed up to the end of Year 6; Q2 refers to exposure in each year of primary education while the number of UA is assessed at the end of the following year; Q3 considers the impact of sustained SEND provision from Year 1 to Year 3, while the outcome is assessed up to the end of Year 6.

#### iii. Causal effects

The typical statistic targeted in randomized controlled trials is the “intention to treat” (ITT) effect, i.e. the population-average impact on the outcome of interest of being randomized to receive or not to receive a particular treatment/intervention. We focussed instead on other effects which are more relevant for policy evaluation, the population average impact of receiving the treatment/intervention (the “average treatment effect”, ATE) and the average impact in the exposed population (the “average treatment effect in the treated”, ATT).

To be able to formally define these causal effects we adopted the concept of potential outcomes (POs) [9], i.e. we considered hypothetical scenarios where the targeted population would, or would not, experience a particular exposure/intervention at a given time point (as for Q1 and Q2) or at multiple time points (as for Q3). Supplementary Box 1 provides full definitions of ATE and ATT in terms of mean POs; these comparisons can be made on the risk differences [RD] or risk ratios [RR] scale. We focussed on both effects for Q1 and Q2 because we were interested in the long- and short-term impacts of SEND provision in the selected populations (children with CLP and with CP with no additional major abnormalities) as well as in the subgroups of those among them who had experienced SEND provision. Effects were calculated for the outcome number of UA standardized by the number of possible sessions, *i.e.* for the rate of UA *(total count of UA)/(maximum count of sessions)*. For Q3 we targeted the ATE of sustained SEND over three years on rates of UA up to the end of primary education.

#### iv. Assumptions

Causal effects are concepts that involve “what if” statements to quantify what would occur under different interventions. To be able to estimate them, different assumptions are invoked. There are several methods available, all of which involve the core assumption that the interventions are well-defined (see Glossary in the Supplementary Material). Further assumptions are invoked by each estimation method. For time-fixed interventions (as those examined by Q1 and Q2), most estimation methods rely on the no unmeasured confounding (NUC) assumption. One can avoid the NUC assumption if an instrumental variable (IV) is available (see Glossary).

#### v. Estimation and interpretation

NUC-based methods for time-fixed interventions (Q1 and Q2) include g-computation and propensity score (PS) methods, such as inverse probability weighting (IPW) and augmented IPW (AIPW) [5, 8] but they differ in terms of which assumptions they invoke in addition to the core assumptions (Supplementary Box 2). In general, IPW (and AIPW) has the advantage of directly identifying violations of the positivity assumption (see Glossary) that may occur when certain covariate–exposure combinations are rare or absent. Wald-type methods such as 2-stage-least-square (2SLS) estimation and generalised methods of moments (GMM) are examples of IV-based methods [5, 9]. These effects are to be interpreted as ATE, ATT, or “local average treatment effect” (LATE) according to which additional assumption is invoked. Generalizations of some of these methods are available to estimate causal contrasts that involve sustained interventions such as for Q3: those mostly used are generalizations of g-computation and IPW [9].

Supplementary Table 1 outlines the five steps we used to address Q1 for the CLP cohort.

### Implementation

#### Simulated Data

Before embarking on analyses of our two populations of interest, we practised estimation and interpretation of results using simulated data inspired by the real data. For simplicity we only simulated data up to Year 4, although the original data went up to Year 6. Figure 2 and Supplementary Table 2 describe how we simulated data for 10,000 children inspired by the empirical data.

**Figure 2.**
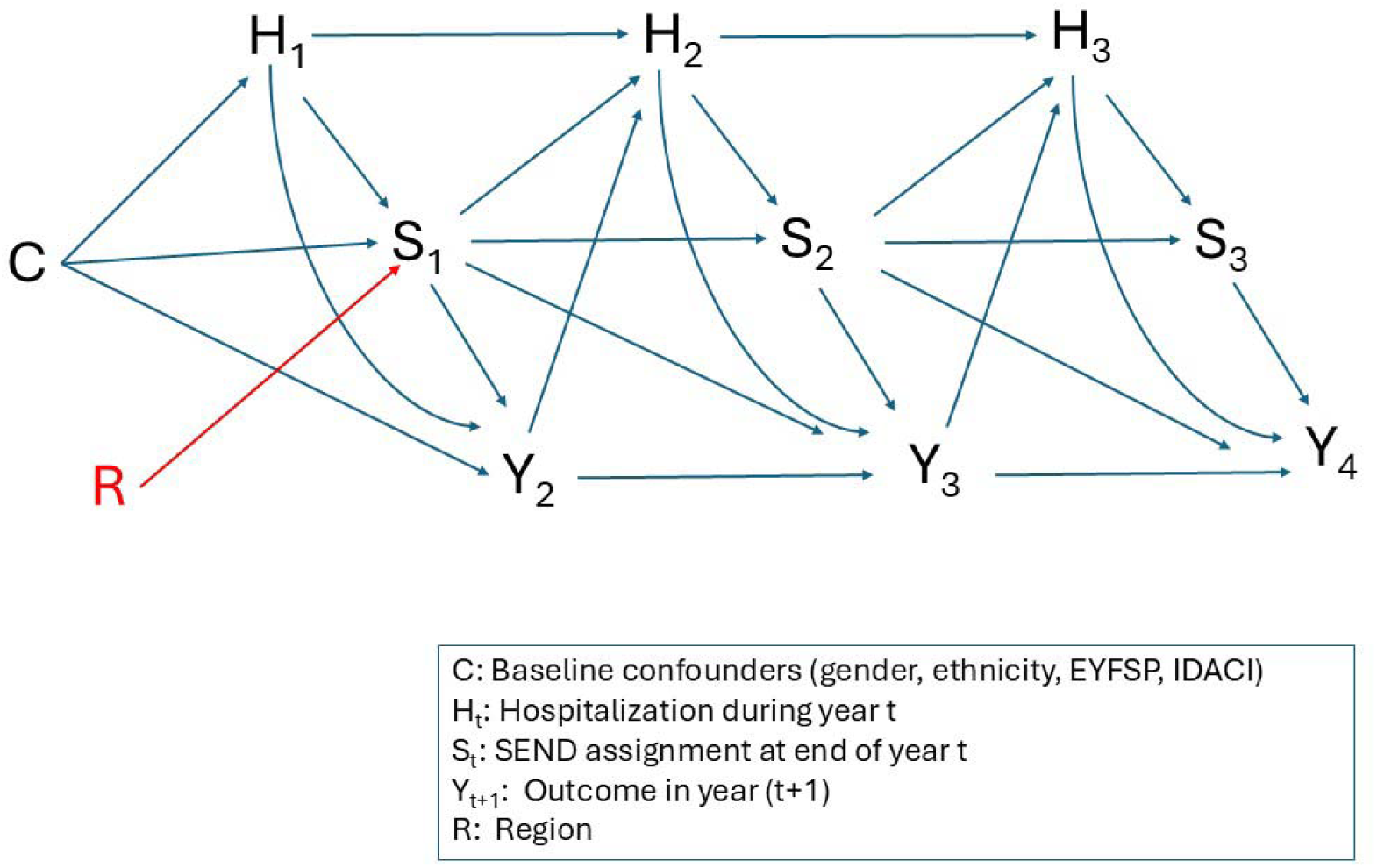
- Directed acyclic graph used to generate the data, with Y being unauthorised absences

In Figure 2, SEND provision is represented by *S*_t_, *t* = 1,2, 3. It im pac ts the outc ome, UA, represented by *Y*_2_, *Y*_3_, *Y*_4_, and simulated as a count variable. *H_t_* is a binary variable representing a time-varying confounder (hospitalization during year t) and *C* represents a set of baseline confounders that influence all other variables. There is also an independent (imaginary) binary variable called “*Region*” which influences only *S*_1_ and thus acts as an IV for *S*_1_ on *Y*. Because we set up the simulated data ourselves, we could calculate the POs corresponding to different interventions (details are in Supplementary Tables 3-5).

#### Estimation methods

We estimated each of the causal contrasts corresponding to Q1-Q3 on the simulated data using three methods that rely on the NUC assumption: g-computation, IPW, and AIPW. When suitable, we also estimated causal contrasts using 2SLS which exploited the simulated IV called Region. For each NUC-based method, we considered departures from its specific assumptions, while including all relevant confounders (thus meeting the NUC assumption). For g-computation, the departures concerned incorrect specifications of the outcome (Y) model; For IPW incorrect specifications of the propensity score (PS) model and for AIPW incorrect specifications of Y and/or PS models. For completeness we considered these effects expressed on both the ratio (i.e. RR) and difference (i.e. RD) scale.

We implemented these methods in Stata and R, with code made available on GitHub (https://github.com/bldestavola/Causal-analyses-using-education-health-linked-data-for-England-a-case-study).

## Results

### Descriptive analysis

The simulated dataset holds information on 10,000 pupils. 2 6.1% of pupils in the sampled data have SEND recorded in Year 1, increasing to 29.4% and 33.2% in Year 2 and 3. The observed UA rates in those not exposed/ exposed in Year 1 are respectively 0.060 and 0.031 (or 6.0 and 3.1 per 100 sessions) and 0.063 and 0.014 in those never/always exposed. Hence, in these simulated data, SEND provision appears to reduce UA rates, but these comparisons may be affected by bias if interpreted causally because of likely unmeasured confounding (“selection bias” in the econometrics literature).

### Causal analysis

***Q1- The long-term effect of SEND:** Does SEND provision in Year 1 affect the number of UA from Year 2 to Year 4?*

Since we know the true population rates under exposure and non-exposure, we also know the true ATE and true ATT (Table 1). On both scales (RR and RD) the true ATT is smaller/more negative than the ATE (on the RR scale: ATT_RR_=29.1% and: ATE_RR_=39.4%; on the RD scale: ATT_RD_=-0.076 and: ATE_RD_=-0.044) indicating the stronger effect of SEND in Year 1 among the treated.

**Table 1.**
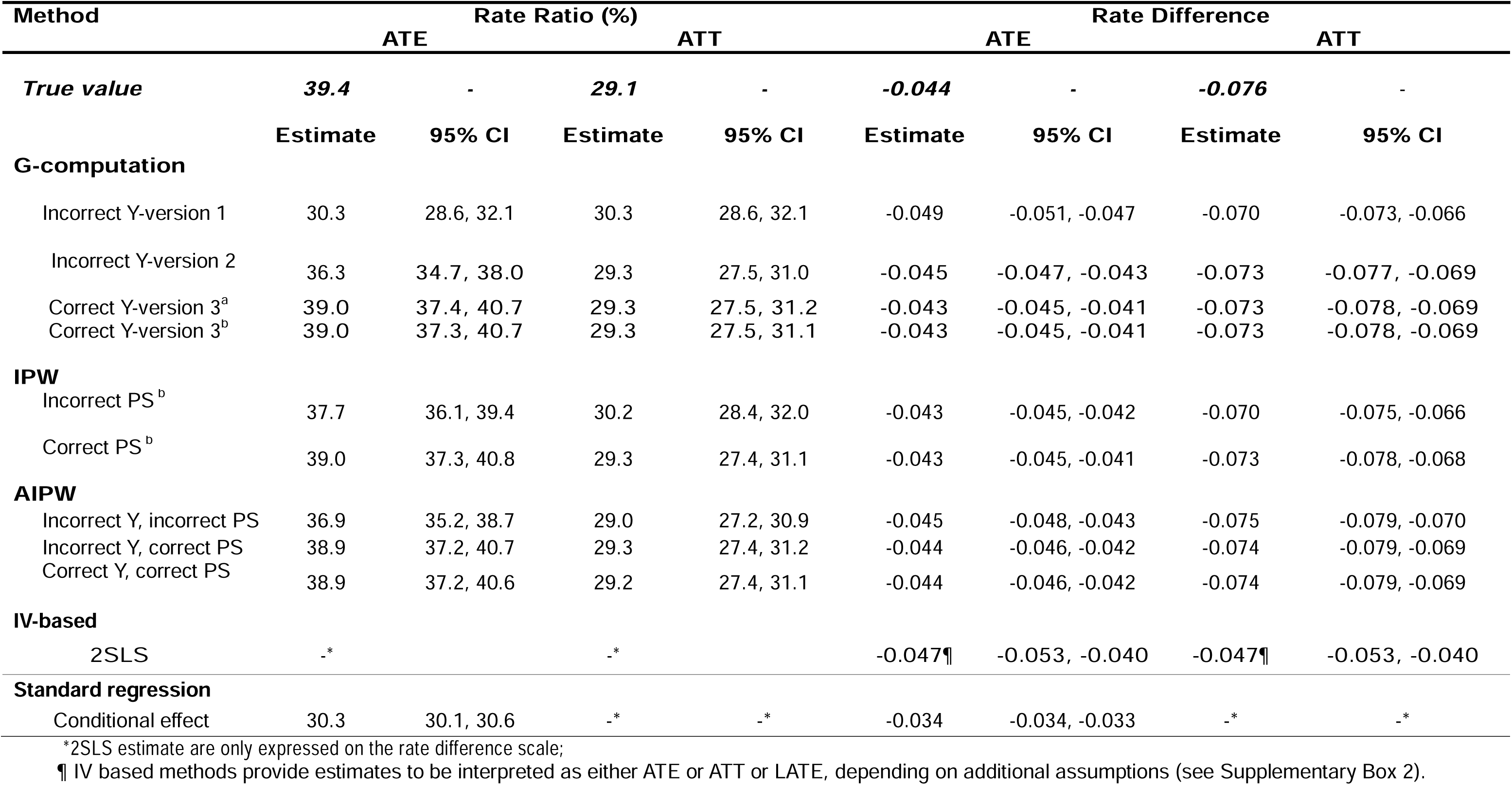
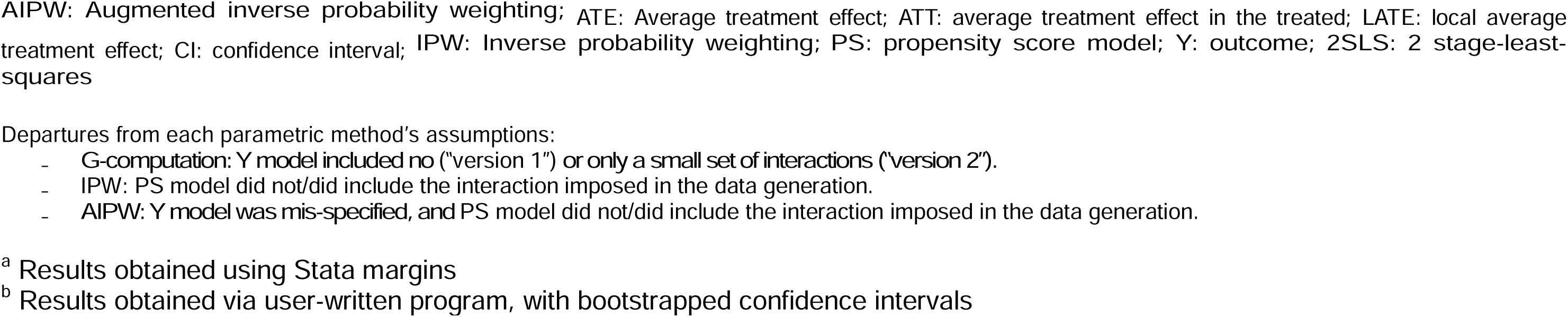
- Addressing Question 1: the long-term effect of SEND provision in Year 1 on cumulative number of unauthorised absences from Year 2 to Year 4, i.e. *S*_1_ on (*Y*_2_ + *Y*_3_ + *Y*_4_). ATE and ATT estimated using alternative methods implemented while meeting or not the methods assumptions; effects are expressed on the rate ratio (RR) and rate difference (RD) scales; HOPE simulated data; N=10,000

| Method | Rate Ratio (%) |  |  |  | Rate Difference |  |  |  |
| --- | --- | --- | --- | --- | --- | --- | --- | --- |
|  | ATE |  | ATT |  | ATE |  | ATT |  |
| **True value** | **39.4** | **-** | **29.1** | **-** | **-0.044** | **-** | **-0.076** | **-** |
|  | **Estimate** | **95% CI** | **Estimate** | **95% CI** | **Estimate** | **95% CI** | **Estimate** | **95% CI** |
| **G-computation** |  |  |  |  |  |  |  |  |
| Incorrect Y-version 1 | 30.3 | 28.6, 32.1 | 30.3 | 28.6, 32.1 | -0.049 | -0.051, -0.047 | -0.070 | -0.073, -0.066 |
| Incorrect Y-version 2 | 36.3 | 34.7, 38.0 | 29.3 | 27.5, 31.0 | -0.045 | -0.047, -0.043 | -0.073 | -0.077, -0.069 |
| Correct Y-version 3<sup>a</sup> | 39.0 | 37.4, 40.7 | 29.3 | 27.5, 31.2 | -0.043 | -0.045, -0.041 | -0.073 | -0.078, -0.069 |
| Correct Y-version 3<sup>b</sup> | 39.0 | 37.3, 40.7 | 29.3 | 27.5, 31.1 | -0.043 | -0.045, -0.041 | -0.073 | -0.078, -0.069 |
| **IPW** |  |  |  |  |  |  |  |  |
| Incorrect PS<sup>b</sup> | 37.7 | 36.1, 39.4 | 30.2 | 28.4, 32.0 | -0.043 | -0.045, -0.042 | -0.070 | -0.075, -0.066 |
| Correct PS<sup>b</sup> | 39.0 | 37.3, 40.8 | 29.3 | 27.4, 31.1 | -0.043 | -0.045, -0.041 | -0.073 | -0.078, -0.068 |
| **AIPW** |  |  |  |  |  |  |  |  |
| Incorrect Y, incorrect PS | 36.9 | 35.2, 38.7 | 29.0 | 27.2, 30.9 | -0.045 | -0.048, -0.043 | -0.075 | -0.079, -0.070 |
| Incorrect Y, correct PS | 38.9 | 37.2, 40.7 | 29.3 | 27.4, 31.2 | -0.044 | -0.046, -0.042 | -0.074 | -0.079, -0.069 |
| Correct Y, correct PS | 38.9 | 37.2, 40.6 | 29.2 | 27.4, 31.1 | -0.044 | -0.046, -0.042 | -0.074 | -0.079, -0.069 |
| **IV-based** |  |  |  |  |  |  |  |  |
| 2SLS | -* |  | -* |  | -0.047¶ | -0.053, -0.040 | -0.047¶ | -0.053, -0.040 |
| **Standard regression** |  |  |  |  |  |  |  |  |
| Conditional effect | 30.3 | 30.1, 30.6 | -* | -* | -0.034 | -0.034, -0.033 | -* | -* |

\*2SLS estimate are only expressed on the rate difference scale;
¶ IV based methods provide estimates to be interpreted as either ATE or ATT or LATE, depending on additional assumptions (see Supplementary Box 2).
AIPW: Augmented inverse probability weighting; ATE: Average treatment effect; ATT: average treatment effect in the treated; LATE: local average treatment effect; CI: confidence interval; IPW: Inverse probability weighting; PS: propensity score model; Y: outcome; 2SLS: 2 stage-least-squares
Departures from each parametric method's assumptions:
- G-computation: Y model included no ("version 1") or only a small set of interactions ("version 2"). - IPW: PS model did not/did include the interaction imposed in the data generation. - AIPW: Y model was mis-specified, and PS model did not/did include the interaction imposed in the data generation.
<sup>a</sup> Results obtained using Stata margins
<sup>b</sup> Results obtained via user-written program, with bootstrapped confidence intervals

We found that, for g-computation, it is crucial that the Y model has a very general specification. Including no interactions with the intervention (SEND in Year 1; see “*incorrect version 1*”), led to estimates that were further away from the true value than an improved version with some interactions (“*incorrect version 2*”; Table 2). However, only when a very general Y model was specified the results were satisfactorily close to the truth. IPW gave estimates which reflected the need for correct PS model specification. It also helped assessing whether the positivity assumption was met (specifically when the PS model was correctly specified). AIPW was confirmed as the most robust approach. As regards the use of *Region* as the IV exploited by 2SLS, we found as expected [13] that estimates were very imprecise, despite *Region* being very strongly associated with the intervention.

**Table 2.** - *Addressing Question 2: the s hort-term effect of SEND, i.e. S*_t_ *on Y_t_*_+1_. ATE and ATT expressed on the rate ratio (RR) scale estimated using correctly specified g-computation; HOPE simulated data, N=10,000.

| Short term effects | Rate Ratio (%) |  |  |  | Rate Difference |  |  |  |
| --- | --- | --- | --- | --- | --- | --- | --- | --- |
|  | ATE |  | ATT |  | ATE |  |  |  |
|  | Estimate | 95% CI | Estimate | 95% CI | Estimate | 95% CI | Estimate | 95% CI |
| $S_1$ on $Y_2$ | | | | | | | | |
| True value | **21.5** |  | **12.7** |  | **-0.046** |  | **-0.077** |  |
| g-computation | 21.5 | 20.8, 22.1 | 13.0 | 12.3, 13.8 | -0.044 | -0.045, -0.044 | -0.074 | -0.077, -0.070 |
| $S_2$ on $Y_3$ | | | | | | | | |
| True value | **17.3** |  | **14.4** |  | **-0.075** |  | **-0.111** |  |
| g-computation | 17.2 | 16.8, 17.6 | 14.2 | 13.5, 15.0 | -0.075 | -0.076, -0.074 | -0.116 | -0.120, -0.111 |
| $S_3$ on $Y_4$ | | | | | | | | |
| True value | **14.3** |  | **13.5** |  | **-0.089** |  | **-0.124** |  |
| g-computation | 14.3 | 14.0, 14.6 | 13.5 | 12.9, 14.1 | -0.088 | -0.089, -0.088 | -0.126 | -0.130, -0.121 |
| Average of $S_t$ on $Y_{(t+1)}$ | | | | | | | | |
| g-computation | 16.9 | 16.7, 17.2 | 15.0 | 14.5, 15.6 | -0.069 | -0.070, -0.069 | -0.104 | -0.107, -0.102 |

ATE: Average treatment effect; ATT: average treatment effect in the treated; CI: confidence interval

For completeness, Table 1 also reports estimated conditional effects of SEND obtained from fitting a Poisson model for the outcome Y with all the confounders but no interactions, as one would normally do in standard regression analyses. Such estimates could have a causal interpretation if the Y model were correctly specified, which clearly it is not.

***Q2 - The short-term effect of SEND:** Does SEND provision in* Year *t* affect the rate of UA in Year *t* + 1?

Table 2 shows the results of sequentia ly estimating the effect of providing SEND one year onto UA rates in the next year. The true ATEs and ATTs indicate that the short-term effects of (simulated) SEND provision increases over time and that it is always stronger in the treated.

Noteworthy is also the relative size of the short- and long-term effects of SEND in Year 1, showing the importance of asking both questions. For example, on the RR scale, the true relative impact in the short-term is to reduce the rate to around 20% while the long-term effect is around 40% (cf. Table 1 and 2).

We used only correctly implemented g-computation to illustrate the closeness of the estimates to the true values; results obtained using IPW and AIPW were very similar but with wider confidence intervals (when implemented correctly). Interestingly, ignoring the trend of these causal associations, and estimating an average short-term effect (last entries in Table 2), as done in dynamic panel data models [14], would miss an important aspect of the causal process. It is recommended therefore to check the assumption of constant effects before drawing any causal conclusions.

**Q3 - Sustained effect:** Does sustained SEND provision in Year 1, 2 and 3 affect the rate of UA up to Year 4?

The true ATEs for sustained exposure to SEND for 3 years on UA rates up to the end of Year 4 is 12.8% on the RR scale and – 0.089 (a decrease of 8.9 absences per 100 sessions) on the RD scale (Table 3). This is a setting affected by time-varying confounding because the impact of sustained SEND is both confounded by contemporaneous H and mediated by later H. Traditional regression analysis that controls for all values of H leads to biased estimates of the causal effect of sustained SEND because it blocks the indirect effect of SEND that works via H and also might introduce collider bias if H and the outcome have common causes [15]. This is indeed what is seen in our results (Table 3). In contrast, both g-computation and IPW- if correctly implemented for sustained interventions- deal with time- varying confounding and l ead to estimates which are closer to the truth. Of the two methods however, IPW, despite being the less precise, is preferable because it relies on fewer modelling assumptions. It only requires the correct specification of the PS models at each intervention point (here Year 1, Year 2 and Year 3), while g-computation relies on the correct specification of the models for time-varying H and for time-varying Y at each time point after baseline (i.e. Year 2, Year 3 and Year 4).

**Table 3.**
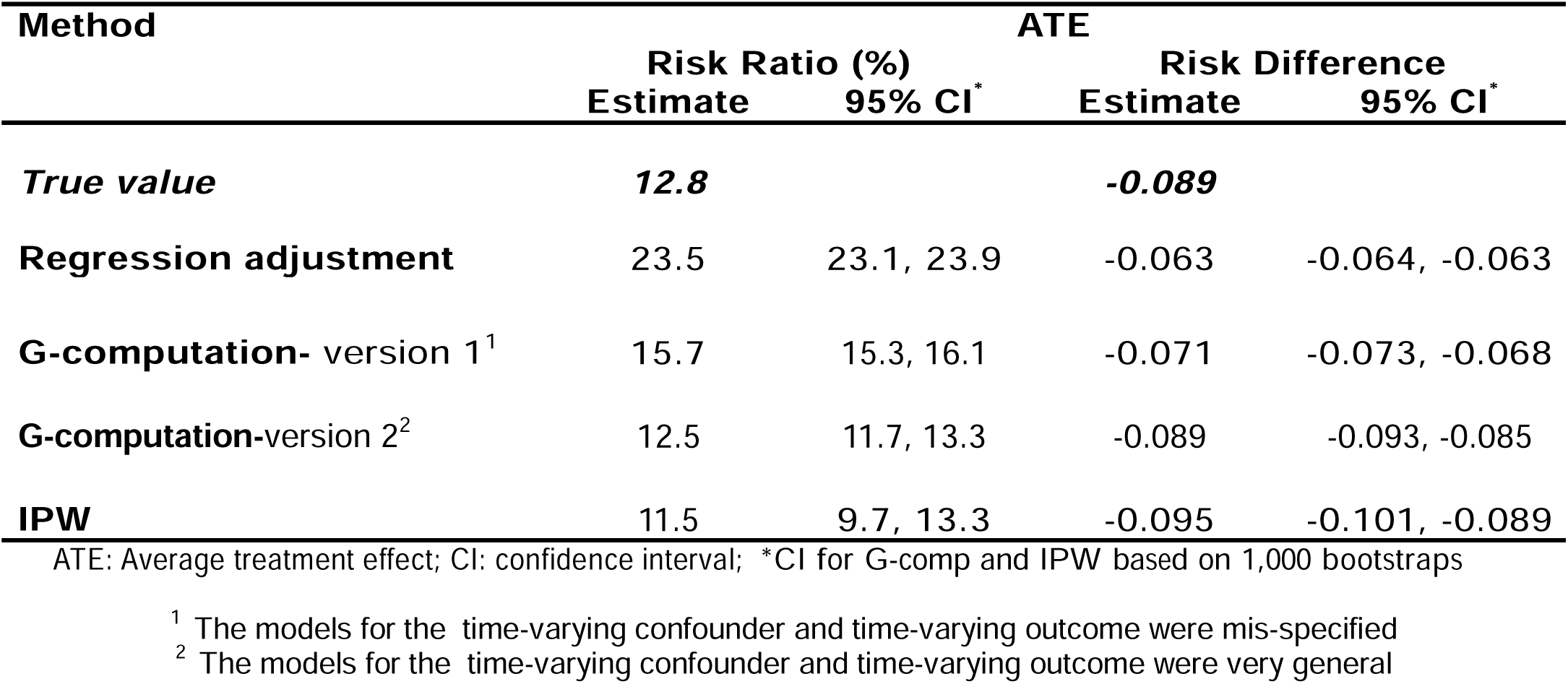
- *Addressing Question 3: the joint effect of* (*S*_1_, *S*_2_*, S*_3_) *on* (*Y*_2_ + *Y*_3_ + *Y*_4_). ATE expressed on the rate ratio (RR) and rate difference (RD) scale estimated using alternative methods; HOPE simulated data; N=10,000.

| Method | ATE |  |  |  |
| --- | --- | --- | --- | --- |
|  | Risk Ratio (%)<br>Estimate | 95% CI* | Risk Difference<br>Estimate | 95% CI* |
| **True value** | **12.8** |  | **-0.089** |  |
| **Regression adjustment** | 23.5 | 23.1, 23.9 | -0.063 | -0.064, -0.063 |
| **G-computation- version 1<sup>1</sup>** | 15.7 | 15.3, 16.1 | -0.071 | -0.073, -0.068 |
| **G-computation-version 2<sup>2</sup>** | 12.5 | 11.7, 13.3 | -0.089 | -0.093, -0.085 |
| **IPW** | 11.5 | 9.7, 13.3 | -0.095 | -0.101, -0.089 |

ATE: Average treatment effect; CI: confidence interval; \*CI for G-comp and IPW based on 1,000 bootstraps
<sup>1</sup> The models for the time-varying confounder and time-varying outcome were mis-specified
<sup>2</sup> The models for the time-varying confounder and time-varying outcome were very general

## Discussion

Administrative data have the potential for addressing questions of policy effectiveness when the target population, intervention and likely confounders are reliably captured in the data. However, researchers may find that such endeavours cannot be attempted with the available data, or that the original questions should be reformulated [16]. In our example the overarching question of SEND effectiveness in the general population had to be restricted to specific clinical populations likely to benefit from SEND provision and to share similar needs. For these populations the mechanisms underlying the non-randomised nature of the intervention were sufficiently known and the available data believed to be sufficiently exhaustive to attempt causal inference. This, however, limits the generalisability of the results to larger populations.

In using simulated data, we demonstrated that implementation of estimation methods involved strong modelling (parametric) assumptions, and that only the specification of very general models led to sensible results, especially when implementing g-computation. We are sharing our code and simulated data so that researchers addressing similar policy questions can apply and compare results from applying these methods. However unverifiable assumptions such as, respectively, NUC and IV need to be critically assessed before attempts at causal inference can be justified.

## Conclusion

We learned that when using administrative data, fully specifying the causal question is an iterative process. We found that we needed to modify our questions according to the available data. Simulated data can help researchers practice implementation and interpretation of their results.

## Acknowledgments

Members of the HOPE study team include: Ruth Gilbert (PI), Katie Harron, Bianca L De Stavola, Lorraine Dearden, Tamsin Ford (senior work package leads), Kate Lewis, Vincent Nguyen, Ania Zylbersztejn, Jennifer Saxton, Jacob Matthews, William Farr (leading roles in the 4 work packages), Ayana Cant, Laura Gimeno, Isaac Winterburn, Andrea Aparicio Castro, Julia Shumway, Lucy Karwatowska (contributing researchers), Matthew Lilliman (programme manager), Kate Boddy (public engagement coordinator), Stuart Logan, Jugnoo Rahi, Kristine Black-Hawkins, Johnny Downs (co-investigators).

We are grateful to the HOPE study Advisory Group: Chris Bonell (chair, professor of health and sociology, London School of Hygiene and Tropical Medicine), Kate Evans-Jones, Julia Ogden (public members), Jo Hutchison (Director of SEND, Education Policy Institute), Karen Horridge (consultant community paediatrician, Sunderland).

We gratefully acknowledge all children and families whose de-identified data were used in this research. We thank Ruth Blackburn, Milagros Ruiz, Matthew Jay, Antony Stone and Farzan Ramzan for ECHILD Database support.

The ECHILD Database uses data from the Department for Education (DfE). The DfE does not accept responsibility for any inferences or conclusions derived by the authors. This work contains statistical data from ONS which is Crown Copyright. The use of the ONS statistical data in this work does not imply the endorsement of the ONS in relation to the interpretation or analysis of the statistical data. This work uses research datasets which may not exactly reproduce National Statistics aggregates. Evidence from this research contributes to the NIHR Children and Families Policy Research Unit but was not commissioned by the NIHR Policy Research Programme.

## Ethics Statement

Permissions to use linked, de-identified data from HES and NPxD were granted by NHS England (DARS-NIC-381972-Q5F0V-v0.5) and the Department for Education (DR200604.02B). Ethical approval for the ECHILD project was granted by the National Research Ethics Service (17/LO/1494), NHS Health Research Authority Research Ethics Committee (20/EE/0180 and 21/SW/0159). Separate ethical approval was not required to use de-identified data for the analyses presented in this paper. ECHILD is available for re-use through the Office for National Statistics Secure Research Service (ONS SRS).

## Data Availability Statement

The ECHILD database is made available for free for approved research based in the UK, via the ONS Secure Research Service. Enquiries to access the ECHILD database can be made by emailing. Researchers will need to be approved and submit a successful application to the ECHILD Data Access Committee and ONS Research Accreditation Panel to access the data, with strict statistical disclosure controls of all outputs of analyses.

## Funding

This project is funded by the National Institute for Health Research (NIHR) under its ‘Programme Grants for Applied Research Programme’ (Grant Reference Number NIHR202025, The HOPE Study). The views expressed are those of the authors and not necessarily those of the NIHR or the Department of Health and Social Care. ECHILD is supported by ADR UK (Administrative Data Research UK), an Economic and Social Research Council (part of UK Research and Innovation) programme (ES/V000977/1, ES/X003663/1, ES/X000427/1).

## Statement of Conflicts of Interest

There are no conflicts.

## Abbreviations

AIPW: Augmented inverse probability weighting;
ATE: Average treatment effect;
ATT: average treatment effect in the treated;
CI: confidence interval;
CLP: left lip and/or palate;
CP: cerebral palsy;
ECHILD: Education and Child Health Insights from Linked Data;
EYFSP: early years foundation stage profile;
GMM: Generalised method of moments;
HOPE: Health Outcomes of young People throughout Education;
IDACI: Income Deprivation Affecting Children Index;
IPW: Inverse probability weighting;
PO: potential outcome;
PS: propensity score model;
SEND: special education need and disabilities;
Y: outcome;
2SLS: 2 stage-least-squares.

## Notes

### Competing Interest Statement

The authors have declared no competing interest.

### Author Declarations

Permissions to use linked, de-identified data from HES and NPD were granted by NHS England (DARS-NIC- 381972-Q5F0V-v0.5) and the Department for Education (DR200604.02B). Ethical approval for the ECHILD project was granted by the National Research Ethics Service (17/LO/1494), NHS Health Research Authority Research Ethics Committee (20/EE/0180 and 21/SW/0159).

