## Supplementary Material for "Causal analyses using education-health linked data for England: a case study"

**Authors**

Bianca L De Stavola, [1*] Andrea Aparicio Castro, [1] Vincent G. Nguyen,[1] Kate Lewis,[1] Lorraine Dearden,[2] Katie Harron,[1] Ania Zylbersztejn,[1] Julia Shumway ,[1] Ruth Gilbert,[1] on behalf of the HOPE study team

**Supplementary Box 1 - Formal definition of causal contrasts**

Formally, let $Y$ be the outcome and $A$ the intervention of interest. With $Y$ a continuous variable and $A$ a binary (0/1) variable, we denote $Y^0$ to be the potential outcome (PO) if $A$ were set to 0 and $Y^1$ the PO if $A$ were set to 1 (assuming no interference, see Glossary in Supplementary Material). We could then express the causal effect of $A$ on $Y$ as a comparison of mean POs when $A = 1$ or $A = 0$ for the entire population of interest. This is the Average Treatment Effect (ATE):

$$ATE = E(Y^1) - E(Y^0),$$

where $E$ stands for expectation (i.e. mean) calculated over the full population.

The average treatment effect in the treated (ATT) instead quantifies the impact of removing $A$ from those who had received it (the "treated"):

$$ATT = E(Y^1|A = 1) - E(Y^0|A = 1),$$

where "|" stands for "conditionally on".

With different types of outcomes, different contrasts may be more suitable. For example, if $Y$ were a count variable and $N$ its denominator, we might prefer to compare potential rates in the full population giving the ATE, either on the rate difference (RD) scale:

$$ATE_{RD} = E\left(\frac{Y^1}{N}\right) - E\left(\frac{Y^0}{N}\right),$$

or on the rate ratio (RR) scale:

$$ATE_{RR} = E\left(\frac{Y^1}{N}\right) \Big/ E\left(\frac{Y^0}{N}\right),$$

with equivalent definitions for the ATTs.

Generalizations of these causal contrasts can be made to settings where the intervention is sustained over a period, with added variations in the comparisons of alternative scenarios that may be of interest. For example, we could compare no interventions, versus an intervention for 1 year, versus an intervention for 2 years, an intervention only in Year 2, etc. In this paper we consider contrasts of POs where the intervention is sustained (or not given) over three time points. In the simple setting with a continuous outcome the ATE on the RD scale is defined as:

$$ATE_{RD} = E(Y^{111}) - E(Y^{000})$$

where $Y^{000}$ and $Y^{111}$ are POs had $A$ been set respectively to 1 and to 0 at each of time 1,2, and 3.

**Supplementary Box 2 – Outline of estimation methods used for the simulated data**

| Methods | Description | Assumptions | | | | | | |
|---|---|---|---|---|---|---|---|---|
| | | Positivity | Correct Y | Correct PS | Core IV | Homogeneity | No effect modification | Monotonicity |
| **For time-fixed (baseline) interventions** | | | | | | | | |
| g-computation | It uses modelling of the outcome as a function of the intervention/exposure and confounders to predict POs which are then averaged (”marginalised”) leading to estimating the causal contrast of interest | | √ | | | | | |
| IPW by PS | It relies on the estimated propensity score (PS), that is the probability of receiving the intervention/exposure, to re-balance unequal distributions of the confounders before fitting an outcome model without covariates. | √ | | √ | | | | |
| AIPW | Augmented IPW: combine the methods above to give double robustness to the estimators (we implemented | √ | either | or | | | | |
| 2SLS | Linear regression model which includes a predicted value of the intervention/exposure, obtained after fitting a linear regression model of the exposure/intervention on the IV (the 2 stages) | | √ | | √ | Either* | Either* | Or* |

**Supplementary Box 2 –continued**

| Methods | Description | Assumptions a | | | |
|---|---|---|---|---|---|
| | | Positivity | Correct Y | Correct L | Correct PS |
| **<u>For sustained interventions</u>** | | | | | |
| g-computation | It uses modelling of the first set of POs to predict the following values of the time-varying confounder L, which are then used to predict the next POs, etc., each model being a function of exposure and time-fixed confounders as well; the final POs which are then averaged (”marginalised”) leading to estimating the causal contrast of interest | | √ | √ | |
| IPW by PS | It relies on the estimated PS for the time-varying intervention, that is the probability of receiving the exposure/intervention at each time; the PSs are then used to re-balance unequal distributions of the time-fixed and time-varying confounders before fitting a model of choice for the POs. | √ | | | √ |

PO: potential outcome; PS: propensity score

*The additional assumption of homogeneity of effects is called up when using IV methods to estimate the ATE; the additional assumption of no effect modification by the instrument is called up when using IV methods to estimate the ATT; the additional assumption of monotonicity of effects is called up when using IV methods to estimate the LATE (Hernán, MA; Robins, JM. Instruments for Causal Inference: An Epidemiologist's Dream?. Epidemiology 17(4): 360-372, July 2006).

**Supplementary Table 1: Implementation of target trial emulation to assess the causal effect of SEN by Year 1 on unauthorised school absences by Year 6 in children with cleft lip and/or palate** (Q1: more details in [1])

| | **Ideal target trial** | **Emulated target trial** |
|---|---|---|
| **Eligibility criteria** | Diagnosed with cleft lip and/or palate but no other major abnormalities prior to Year 1<br>Born in England and started Year 1 between 2008/2009 and 2018/2019 | Has a birth and school records in ECHILD<br>Identified in HES with cleft lip and/or palate but no other major abnormalities before start of Year 1 in ECHILD<br>Started Year 1 between 2008/2009 and 2018/2019 |
| **Recruitment period** | Started Year 1 in between the academic years 2008/2009 and 2018/2019 | Started Year 1 between the academic years 2008/2009 and 2018/2019 |
| **Follow-up duration** | From randomization to the intervention to the earliest of:<br>- the end of primary school<br>- loss of follow-up<br>- death<br>- end of study | From January School Census in Year 1 to the earliest of:<br>- the end of primary school<br>- loss of follow-up in ECHILD<br>- death<br>- 31 August 2019 |
| **Outcome** | Unauthorised school absences as defined using half-days | Unauthorised school absences as defined using half days |
| **Interventions to be compared** | SEND provision to be randomly allocated following randomization in Year 1 | SEND provision allocated by the January School Census in Year 1 |
| **Causal contrasts** | The average treatment effect (ATE) and average treatment effect in the treated (ATT) of initiating SEND versus non-initiating SEND by Year 1 on the rate of unauthorised absences, both expressed as rate ratios | Same as for the ideal target trial |
| **Analysis plan** | Negative Binomial Regression of the number of events accountings for duration of follow-up<br><br>Clustering by school and/or local authority to be dealt via robust inference | Appropriate methods for confounding adjustment (regression adjustment, g-computation, IPW and AIPW) for the same outcome models as for the target trial.<br><br>Clustering by school and/or local authority to be dealt via robust inference |

**Supplementary Table 2: Summary of data generating mechanism for the simulated data**

| | |
|---|---|
| **Number of individuals:** | 10,000 |
| **Baseline covariates** | |
| Region A | $R \sim \text{Bernoulli}(0.30)$ |
| Male gender | $C_1 \sim \text{Bernoulli}(0.52)$ |
| White ethnicity | $C_2 \sim \text{Bernoulli}(0.40)$ |
| IDACI score | $C_3 \sim \text{Normal}(\mu_3, 1)$<br>$\mu_3 = 0 - 1 \times I_{\text{White}}$ |
| Poverty | lowest two quintiles of IDACI |
| EYFSP | $C_4 \sim \text{Normal}(\mu_4, 0.5)$<br>$\mu_4 = 0.1 + 0.1 \times I_{\text{White}} - 0.2 \times I_{\text{Male}} - 0.15 \times I_{\text{Poverty}}$ |
| Below average EYFSP | EYFSP $< 0$ |
| Hospitalization at time 1 | $\mathtt{H}_1 \sim \text{Bernoulli}(h_1)$,<br>$logit(h_1) = -3.5 + 0.5 \times I_{\text{Male}} - 0.1 \times I_{\text{White}} + 1 \times I_{\text{Poverty}}$ |
| SEND at time 1 | $\mathtt{S}_1 \sim \text{Bernoulli}(s_1)$,<br>$logit(s_1) = -2.5 + 0.3 \times I_{\text{Male}} - 0.1 \times I_{\text{White}} + 1 \times I_{\text{EYFSP}<0} +$<br>$1 \times I_{\text{Poverty}} + 1 \times H_1 + 2 \times I_{\text{Region}=1} + 1.5 \times I_{\text{EYFSP}<0} \times I_{\text{Poverty}}$ |
| **Number of sessions** | $integer(N_t)$ where $N_t \sim \text{Normal}(\eta_t, 30)$<br>$\eta_t = 360 - 10 \times I_{\text{Poverty}}$ for $t = 2, \ldots, 4$ |
| **Time-varying covariates and outcomes, $t = 2, \ldots, 4$** | |
| Outcome | $Y_t \sim \text{Poisson}(\lambda_t)$<br>$log(\lambda_t) = 8 + 0.25 \times I_{\text{Male}} + 0.5 \times I_{\text{Poverty}} + 0.5 \times I_{\text{EYFSP}<0} +$<br>$1.5 \times H_{t-1} + S_{t-1} \times (-0.5 - 2 \times I_{\text{EYFSP}<0} - 0.2 \times (t-1)) - \ln(\text{den}_t)$ |
| Hospitalization at time $t$ | $\mathtt{H}_t \sim \text{Bernoulli}(h_t)$,<br>$logit(h_t) = -3.5 + 0.5 \times I_{\text{Male}} - 0.1 \times I_{\text{White}} + 1 \times I_{\text{Poverty}} +$<br>$0.5 \times H_{t-1} + 0.5 \times S_{t-1} + 0.1 \times Y_t$ |
| SEND at time $t$ | $\mathtt{S}_t \sim \text{Bernoulli}(s_t)$,<br>$logit(s_t) = -2.5 + 0.3 \times I_{\text{Male}} - 0.1 \times I_{\text{White}} + 1 \times I_{\text{Poverty}} +$<br>$1 \times H_t + 2.5 \times S_{t-1}$ |

**Supplementary Table 2 (cont'd)**

The directions of association are as follows:

Hospitalization (H) depends on:
being male (positively); being white (negatively); being in the bottom 2 quintiles of the IDACI distribution (positively); on having been hospitalised in the previous year (positively) on having SEND recorded in the previous year (positively); and on previous value of Y (positively).

- SEND (S) depends on:
being male (positively); being white (negatively); being in the bottom 2 quintiles of the IDACI distribution (positively); on having a below average EYFSP score (positively); on having been hospitalised in the previous year (positively) and on having SEND recorded in the previous year (positively). In Year 1 SEND also depends on Region.

- Unauthorised absences (Y) depends on: being male (positively); being white (negatively); being in the bottom 2 quintiles of the IDACI distribution (positively); on having being hospitalised in the previous year (positively); on having a below average EYFSP score (positively); and on having SEND recorded in the previous year (negatively), with this effect being modified by EYFSP and by time (increasing with worse EYFSP and with time, see details below).

- The number of sessions depends on whether the child lives in an area in the lowest 2 quintiles of the IDACI distribution (negatively).

- The short-term effect of SEND (i.e. from $S_t$ to $Y_{t+1}$) is set to increase if the child had a below average EYFSP score, and to increase with time *t*.

**Supplementary Table 3: Potential outcomes, confounders and exposures for interventions on S at time 1; HOPE simulated data**

**Potential outcomes setting $S_1 = a$**

$Y_2^a \sim \text{Poisson}(\lambda_2^a)$

$$log(\lambda_2^a) = 8 + 0.25 \times I_{\text{Male}} + 0.5 \times I_{\text{Poverty}} + 0.5 \times I_{\text{EYFSP}<0} + 1.5 \times H_1 + a \times (-0.5 - 2 \times I_{\text{EYFSP}<0} - 0.2 \times 1) - \ln(\text{den}_2)$$

$Y_3^a \sim \text{Poisson}(\lambda_3^a)$

$$log(\lambda_3^a) = 8 + 0.25 \times I_{\text{Male}} + 0.5 \times I_{\text{Poverty}} + 0.5 \times I_{\text{EYFSP}<0} + 1.5 \times H_2^a + S_2^a \times (-0.5 - 2 \times I_{\text{EYFSP}<0} - 0.2 \times 2) - \ln(\text{den}_3)$$

*where:*

$$\text{H}_2^a \sim \text{Bernoulli}(h_2^a),$$
$$logit(h_2^a) = -3.5 + 0.5 \times I_{\text{Male}} - 0.1 \times I_{\text{White}} + 1 \times I_{\text{Poverty}} + 0.5 \times H_1 + 0.5 \times a + 0.1 \times Y_2^a$$

$$\text{S}_2^a \sim \text{Bernoulli}(s_2^a),$$
$$logit(s_2^a) = -2.5 + 0.3 \times I_{\text{Male}} - 0.1 \times I_{\text{White}} + 1 \times I_{\text{Poverty}} + 1 \times H_2^a + 2.5 \times a$$

$Y_4^a \sim \text{Poisson}(\lambda_4^a)$

$$log(\lambda_4^a) = 8 + 0.25 \times I_{\text{Male}} + 0.5 \times I_{\text{Poverty}} + 0.5 \times I_{\text{EYFSP}<0} + 1.5 \times H_3^a + S_3^a \times (-0.5 - 2 \times I_{\text{EYFSP}<0} - 0.2 \times 3) - \ln(\text{den}_4)$$

*where:*

$$\text{H}_3^a \sim \text{Bernoulli}(h_3^a),$$
$$logit(h_2^a) = -3.5 + 0.5 \times I_{\text{Male}} - 0.1 \times I_{\text{White}} + 1 \times I_{\text{Poverty}} + 0.5 \times H_2^a + 0.5 \times S_2^a + 0.1 \times Y_3^a$$

$$\text{S}_3^a \sim \text{Bernoulli}(s_3^a),$$
$$logit(s_3^a) = -2.5 + 0.3 \times I_{\text{Male}} - 0.1 \times I_{\text{White}} + 1 \times I_{\text{Poverty}} + 1 \times H_3^a + 2.5 \times S_2^a$$

$S_t^a, H_t^a, Y_t^a$: potential exposure, confounder and outcome at times $t$ following intervention on $S_1$

**Supplementary Table 4: Potential outcomes, time-varying confounders and exposures for repeated interventions on S; HOPE simulated data**

**Potential outcomes setting** $S_1 = a, S_2 = a, S_3 = a$

$Y_2^a \sim \text{Poisson}(\lambda_2^a)$

$$log(\lambda_2^a) = 8 + 0.25 \times I_{\text{Male}} + 0.5 \times I_{\text{Poverty}} + 0.5 \times I_{\text{EYFSP}<0} + 1.5 \times H_1 + a \times (-0.5 - 2 \times I_{\text{EYFSP}<0} - 0.2 \times 1) - \ln(\text{den}_2)$$

$Y_3^{aa} \sim \text{Poisson}(\lambda_3^{aa})$

$$log(\lambda_3^{aa}) = 8 + 0.25 \times I_{\text{Male}} + 0.5 \times I_{\text{Poverty}} + 0.5 \times I_{\text{EYFSP}<0} + 1.5 \times H_2^a + a \times (-0.5 - 2 \times I_{\text{EYFSP}<0} - 0.2 \times 2) - \ln(\text{den}_3)$$

*where:*

$$\text{H}_2^a \sim \text{Bernoulli}(h_2^a),$$
$$logit(h_2^a) = -3.5 + 0.5 \times I_{\text{Male}} - 0.1 \times I_{\text{White}} + 1 \times I_{\text{Poverty}} + 0.5 \times H_1 + 0.5 \times a$$

$Y_4^{aaa} \sim \text{Poisson}(\lambda_4^{aaa})$

$$log(\lambda_4^{aaa}) = 8 + 0.25 \times I_{\text{Male}} + 0.5 \times I_{\text{Poverty}} + 0.5 \times I_{\text{EYFSP}<0} + 1.5 \times H_3^{aa} + a \times (-0.5 - 2 \times I_{\text{EYFSP}<0} - 0.2 \times 3) - \ln(\text{den}_4)$$

*where:*

$$\text{H}_3^{aa} \sim \text{Bernoulli}(h_3^{aa}),$$
$$logit(h_3^{aa}) = -3.5 + 0.5 \times I_{\text{Male}} - 0.1 \times I_{\text{White}} + 1 \times I_{\text{Poverty}} + 0.5 \times H_2^a + 0.5 \times a$$

$H_1^a, Y_1^a$: potential time-varying confounder and outcome at times 2 following intervention on $S_1$;
$H_3^{aa}, Y_3^{aa}$: potential time-varying confounder and outcome at times 2 following intervention on $S_1, S_2$;
$Y_4^{aaa}$: potential time-varying confounder and outcome at times 2 following intervention on $S_1, S_2, S_3$.

### Supplementary Table 5: Data Structure

The data are longitudinal and start by keeping then in long format. For example, the first five individuals have these data:

| id | t | region | male | white | idaci | eyfsp | Hosp | SEN | Y | den |
|---|---|---|---|---|---|---|---|---|---|---|
| 2 | 1 | 1 | 1 | 1 | -1.597499 | -.3910303 | 0 | 1 | . | 321 |
| 2 | 2 | 1 | 1 | 1 | -1.597499 | -.3910303 | 0 | 0 | 1 | 366 |
| 2 | 3 | 1 | 1 | 1 | -1.597499 | -.3910303 | 0 | 0 | 20 | 397 |
| 2 | 4 | 1 | 1 | 1 | -1.597499 | -.3910303 | 0 | 0 | 23 | 354 |
| 191 | 1 | 0 | 0 | 1 | -.887997 | .2439614 | 1 | 0 | . | 359 |
| 191 | 2 | 0 | 0 | 1 | -.887997 | .2439614 | 1 | 0 | 39 | 362 |
| 191 | 3 | 0 | 0 | 1 | -.887997 | .2439614 | 1 | 0 | 43 | 344 |
| 191 | 4 | 0 | 0 | 1 | -.887997 | .2439614 | 0 | 0 | 38 | 320 |
| 194 | 1 | 0 | 1 | 0 | .157719 | .3670338 | 0 | 0 | . | 364 |
| 194 | 2 | 0 | 1 | 0 | .157719 | .3670338 | 1 | 0 | 27 | 274 |
| 194 | 3 | 0 | 1 | 0 | .157719 | .3670338 | 1 | 0 | 80 | 336 |
| 194 | 4 | 0 | 1 | 0 | .157719 | .3670338 | 1 | 1 | 75 | 364 |
| 299 | 1 | 1 | 0 | 0 | -2.952704 | .2799487 | 0 | 1 | . | 341 |
| 299 | 2 | 1 | 0 | 0 | -2.952704 | .2799487 | 0 | 1 | 4 | 395 |
| 299 | 3 | 1 | 0 | 0 | -2.952704 | .2799487 | 0 | 1 | 5 | 366 |
| 299 | 4 | 1 | 0 | 0 | -2.952704 | .2799487 | 0 | 1 | 3 | 368 |
| 317 | 1 | 1 | 0 | 0 | -.1047611 | .1998861 | 0 | 0 | . | 347 |
| 317 | 2 | 1 | 0 | 0 | -.1047611 | .1998861 | 0 | 0 | 12 | 373 |
| 317 | 3 | 1 | 0 | 0 | -.1047611 | .1998861 | 0 | 0 | 13 | 372 |
| 317 | 4 | 1 | 0 | 0 | -.1047611 | .1998861 | 0 | 0 | 11 | 378 |

The `id` are not consecutive because the dataset is a random sample taken from a population of 1,00,000. The outcome $Y$ is defined only from time 2; only `Hosp`, `SEN`, `Y` and `den` vary with time.
The same information could be view in wide format (the list does not include the time fixed covariates in order to keep one row per individual):

| id | Hosp1 | SEN1 | Y2 | den2 | Hosp2 | SEN2 | Y3 | den3 | Hosp3 | SEN3 | Y4 |
|---|---|---|---|---|---|---|---|---|---|---|---|
| 2 | 0 | 1 | 1 | 366 | 0 | 0 | 20 | 397 | 0 | 0 | 23 |
| 191 | 1 | 0 | 39 | 362 | 1 | 0 | 43 | 344 | 1 | 0 | 38 |
| 194 | 0 | 0 | 27 | 274 | 1 | 0 | 80 | 336 | 1 | 0 | 75 |
| 299 | 0 | 1 | 4 | 395 | 0 | 1 | 5 | 366 | 0 | 1 | 3 |
| 317 | 0 | 0 | 12 | 373 | 0 | 0 | 13 | 372 | 0 | 0 | 11 |

**Glossary**

| Term | Description |
|---|---|
| **No interference** | This assumption is typically invoked to justify the concept of potential outcome**s**. The assumption states that the (distribution of) potential outcomes under different possible interventions for a given individual is the response to a change specific to that individual and is independent of interventions received by others. |
| **Causal consistency** | This states that the potential outcome of an intervention setting the intervention to take the value that is actually observed is equal to the observed outcome, i.e. the potential outcome under observed exposure is *factual* and observed. For counterfactual consistency to hold, the exposure should be "well-defined" in the sense that, if there were different ways of setting it , they would all result in the same potential outcome ("treatment variation invariance"). |
| **Confounding** | The bias caused by shared causes of intervention and outcome. This can be removed by adjustment. This is referred to as "selection bias" in the econometrics literature. |
| **Positivity** | An assumption often made in causal inference, stating that for all individuals in the population of interest, the (conditional) probability to be assigned to any of the intervention groups is larger than 0. |
| **No unmeasured confounding** | The assumption that there all confounding affecting the association between intervention and outcome is accounted for (e.g. by adjustment). |
| **Instrumental variable (IV)** | A variable that is associated with the intervention but does not have a direct association with the outcome of interest.<br>The defining properties of an IV are: (1) the IV must be associated with the intervention, (2) it must be independent of any unmeasured factors U confounding the intervention/outcome relation, and (3) it must be conditionally independent of the outcome given intervention and unmeasured confounders. |
| **Target trial emulation (TTE)** | A framework that applies the study design principles of randomised trials to observational studies: it aims to estimate the causal effect of an intervention. This approach aims to reduce bias and strengthen causal validity in observational studies. |
